# What Constitutes High Risk for Venous Thromboembolism? Comparing Approaches to Determining an Appropriate Threshold

**DOI:** 10.1101/2024.08.30.24312871

**Authors:** Benjamin G Mittman, Bo Hu, Rebecca Schulte, Phuc Le, Matthew A Pappas, Aaron Hamilton, Michael B Rothberg

## Abstract

**Background:** Guidelines recommend pharmacological venous thromboembolism (VTE) prophylaxis only for high-risk patients, but the probability of VTE considered “high-risk” is not specified. Our objective was to define an appropriate probability threshold (or range) for VTE risk stratification and corresponding prophylaxis in medical inpatients.

**Methods:** Patients were adults admitted to any of 10 Cleveland Clinic Health System hospitals between December 2020 and August 2021 (N = 41,036). Hospital medicine physicians and internal medicine residents from included hospitals were surveyed between June and November 2023 (N = 214). We compared five approaches to determining a threshold: decision analysis, maximizing the sensitivity and specificity of a logistic regression model, deriving a probability from a point-based model, surveying physicians’ understanding of VTE risk, and deriving a probability from physician behavior. For each approach, we determined the probability threshold above which a patient would be considered high-risk for VTE. We applied each threshold to the Cleveland Clinic VTE risk assessment model (CCM) and calculated the percentage of the 41,036 patients in our cohort who would be considered eligible for prophylaxis due to their high-risk status. We compared these hypothetical prophylaxis rates with physicians’ observed prophylaxis rates.

**Results:** The different approaches yielded thresholds ranging from 0.3% to 5.4%, corresponding inversely with hypothetical prophylaxis rates of 0.2% to 75%. Multiple thresholds clustered between 0.52% to 0.55%, suggesting an average hypothetical prophylaxis rate of approximately 30%, whereas physicians’ observed prophylaxis rates ranged from 48% to 76%.

**Conclusions:** Multiple approaches to determining a probability threshold for VTE prophylaxis converged to suggest an optimal threshold of approximately 0.5%. Other approaches yielded extreme thresholds that are unrealistic for clinical practice. Physicians prescribed prophylaxis much more frequently than the suggested rate of 30%, indicating opportunity to reduce unnecessary prophylaxis. To aid in these efforts, guidelines should explicitly quantify high-risk.

## Introduction

Venous thromboembolism (VTE) affects 300,000 to 600,000 people and causes up to 100,000 deaths each year in the United States (US), with at least half of all cases attributable to current or recent hospitalization.^1–4^ Multiple randomized controlled trials in medical inpatients have demonstrated reduced rates of symptomatic VTE with low molecular weight heparin (LMWH) prophylaxis, compared to placebo.^5–8^ However, LMWH increases rates of heparin-induced thrombocytopenia (HIT) and bleeding,^9,10^ rendering indiscriminate use harmful and expensive. Therefore, the American College of Chest Physicians (ACCP),^11^ American Society for Hematology (ASH),^12^ American Heart Association (AHA),^13^ International Society of Thrombosis and Haemostasis (ISTH),^13^ and American College of Physicians (ACP)^14^ all recommend pharmacological prophylaxis for medical inpatients only if they are at high risk for VTE.

However, none of these guidelines define the *probability* of VTE that should be considered high-risk. Instead, high-risk patients are described as a category based on particular risk factors or settings of care. Some guidelines suggest risk prediction scoring systems, but these point-based scoring systems do not quantify the probability of VTE.^15,16^ This lack of clarity may promote clinician-level variation in the use of VTE prophylaxis and contribute to overuse. Clinicians’ understanding of risk factors and prescription of pharmacological prophylaxis vary substantially, leading to variation in practice.^17^ Establishing an accepted probability threshold for prophylaxis could help to reduce variation and improve quality of care.

There are several ways that a probability threshold could be determined, including theoretical and empirical methods. We compared five distinct approaches to deriving a threshold and compared the resulting thresholds in terms of the percentage of patients who would be considered high-risk and thus potentially eligible for pharmacological prophylaxis. We then compared these percentages to the observed percentage of patients who received prophylaxis.

## Methods

### Setting and Participants

We used five distinct approaches to derive probability thresholds or ranges. These approaches used one physician sample and one patient sample, each drawn from 10 hospitals of the Cleveland Clinic Health System. Hospitals were located in Ohio and Florida and varied in size from a 126-bed community hospital to a 1,400-bed quaternary care academic medical center. The physician sample included internal medicine residents and attending hospitalists who were surveyed between June and November 2023. The patient sample consisted of adult medical patients ≥18 years of age admitted to any Cleveland Clinic hospital between December 2020 and August 2021, excluding surgical, intensive care unit, or COVID-19 positive patients. All patient data were extracted from the Cleveland Clinic electronic health record (EHR) system and verified for accuracy and completeness. The study was approved by the Cleveland Clinic Institutional Review Board (IRBs #22-321 and #14-240).

### Approach 1: Decision Analysis

Decision analytic models compare the expected value of a decision—in this case prescribing LMWH for VTE prophylaxis or not—based on the expected outcomes. Each outcome was valued in terms of cost and utility and the outcomes weighted based on their probability of occurring. The model has been published previously.^18^ We selected two thresholds. The first was the value at which the cost-effectiveness of prophylaxis was exactly $100,000/quality-adjusted life year (QALY), because probabilities of VTE above that threshold would be “cost-effective” based on a generally accepted willingness-to-pay. Thus, high-risk patients would be those for whom prophylaxis was cost-effective. The second threshold represented the point at which the expected value of prophylaxis in QALYs exactly equaled the expected value of no prophylaxis. In this scenario, high-risk patients are those who are expected to derive any net benefit from prophylaxis, regardless of the cost.

### Approach 2: Maximize Sensitivity and Specificity of a Logistic Regression Model

The Cleveland Clinic Model (CCM) is a validated prediction model that computes personalized VTE risk in medical patients based on the most important risk factors.^19^ The model was developed in a sample of approximately 155,000 patients at 10 Cleveland Clinic hospitals in Ohio and Florida and has been externally validated in a separate sample of Michigan hospitals. The Youden Index, derived from the receiver operating characteristics (ROC) curve, summarizes the overall accuracy of a prediction model and identifies a threshold value that maximizes the sum of sensitivity and specificity. We selected the probability threshold that maximized the Youden Index of this prediction model.

### Approach 3: Derive a Probability from a Point-Based Model

The Padua score is a validated risk assessment model derived from medical inpatients in Padua, Italy.^16^ The Padua score is calculated by assigning point values for different risk factors and summing them. A score of four or more is considered high-risk by current guidelines. To convert this score into a probability, we used the CCM to calculate the risk of all patients in our sample with a Padua score of four. We report the mean, median, and range of probabilities for these patients.

### Approach 4: Survey Physicians

We elicited physicians’ stated threshold directly via survey. We asked two questions in the context of medical inpatients: (1) “What probability of developing VTE during hospitalization would you consider high-risk?” and (2) “What is the largest number of patients that you would be willing to give prophylaxis to in order to prevent one VTE?” Question 1 (Q1) included a slider from 0-10% and question 2 (Q2) was free text response. Q1 assessed physicians’ threshold directly, whereas Q2 assessed it indirectly.

In Q2, we excluded blank responses, text answers that did not correspond with a number, and values less than one, which suggested the question was misunderstood. We also excluded two overly influential values (two thousand and one million) identified as outliers. For each question we computed the mean, median, and range of eligible values. To calculate a threshold for Q2, we divided one by the number to treat and then multiplied the result by 37%, which is the estimated efficacy of LMWH in preventing VTE based on a meta-analysis.^20,21^ We used Pearson’s correlation to measure the degree of correspondence between the thresholds derived from the two questions.

### Approach 5: Examine Physician Behavior

We measured physicians’ prophylaxis rates among medical inpatients at Cleveland Clinic hospitals during a period in which prophylaxis was guided by mandatory use of CCM as an EHR-embedded risk calculator. Some physicians nevertheless declined decision support and were able to order prophylaxis without first calculating a patient’s estimated risk. We compared prophylaxis rates between the calculator-guided and nonguided groups.

To determine the implied threshold based on physician behavior, we identified the predicted risk at which 50% of patients received prophylaxis, suggesting equipoise regarding the benefits and harms of prophylaxis. We did this by fitting a simple logistic regression model with the CCM predicted risk as the independent variable and prophylaxis receipt as the outcome. We then used the model coefficients to calculate the predicted risk that corresponded with a 50% probability of receiving prophylaxis.

### Risk Stratification Based on the Different Thresholds

For each threshold produced by the different approaches, we calculated the percentage of patients in our cohort that would be considered high-risk based on predicted probability of VTE from the CCM. We plotted thresholds versus percent of high-risk patients (i.e., those potentially eligible for prophylaxis) and compared them to the observed prophylaxis rates during our study period.

## Results

### Physician & Patient Samples

A total of 224 out of 434 physicians who were contacted completed the survey for a response rate of 51.6%. After excluding ten (4.5%) physicians who failed to answer Q1 or provided an ineligible response to Q2, our final sample contained 214 physicians. For the patient cohort, we identified 43,242 adult medical inpatients that met the eligibility criteria. After removing 2,206 (5.1%) patients with missing data, the final patient cohort contained 41,036 patients, of whom 35,442 (86.4%) had a physician who used the embedded risk calculator.

### Approach 1: Decision Analysis

In the cost-effectiveness analysis, prophylaxis had an incremental cost-effectiveness ratio of $100,000/QALY at a probability of VTE of 1.0% or greater. Ignoring costs, patients had a net benefit from prophylaxis if the probability of VTE was at least 0.3%. Using the 1.0% threshold, 7.0% of inpatients would be considered high-risk, versus 75.4% for the 0.3% threshold (Figure 1).

**Figure 1.**
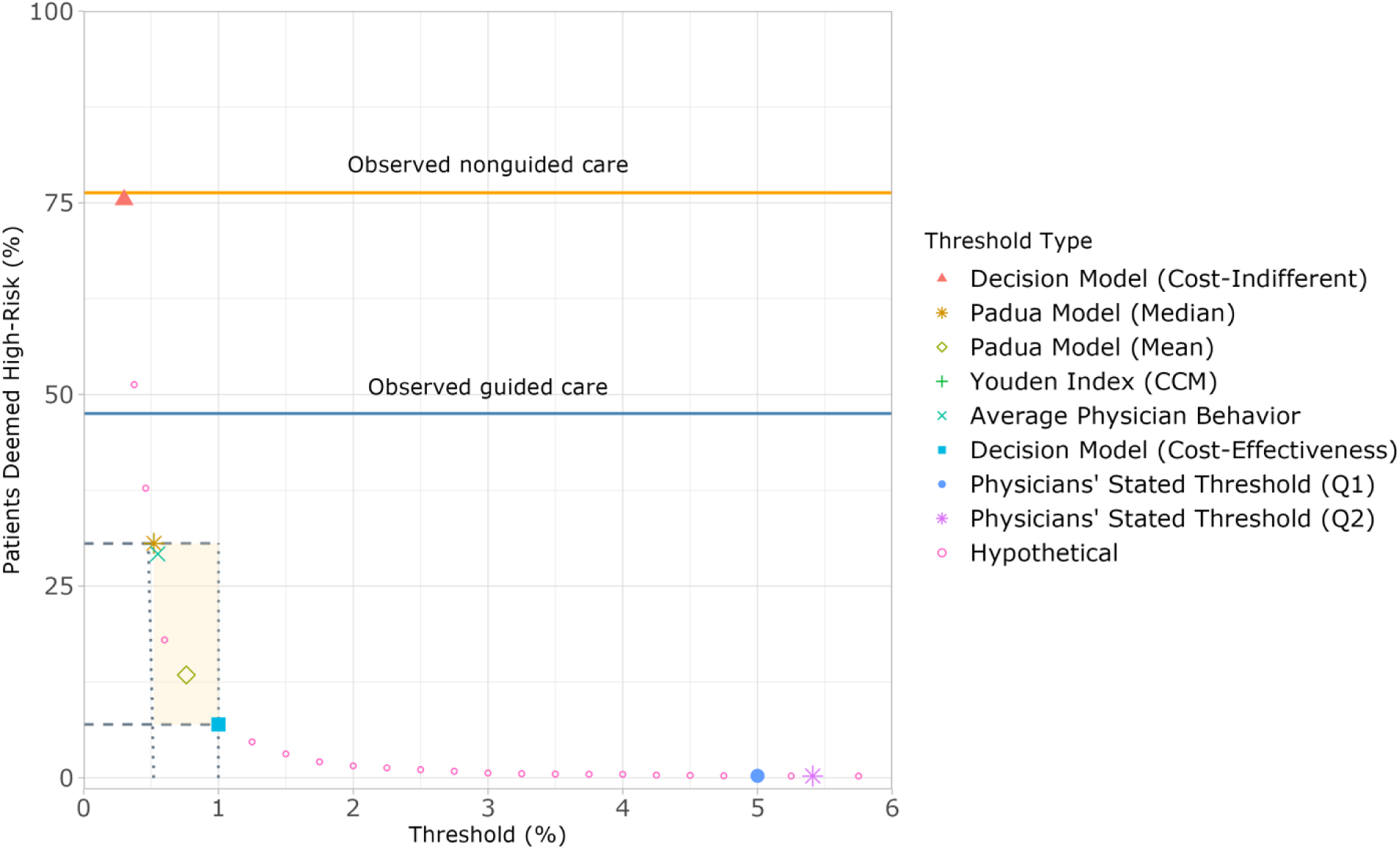
Empirical and theoretical VTE risk thresholds versus the percentage of patients deemed high-risk. The orange and blue horizontal lines indicate observed average prophylaxis rates for all patients who received nonguided and guided care, respectively. The ideal threshold likely falls between 0.52% and 1.0%, with corresponding high-risk percentages ranging between approximately 7% and 31%, represented by the intersecting gray lines and shaded box. Note: Padua Model (Median) and Youden Index (CCM) data points overlap completely because they yielded identical values.

### Approach 2: Maximize Sensitivity and Specificity of a Logistic Regression Model

The threshold based on the Youden Index was 0.52% for the CCM.^19^ At this threshold, 30.6% of patients would be high-risk.

### Approach 3: Derive a Probability from a Point-Based Model

A total of 3,151 (7.7%) patients had a Padua score of exactly four. For them, the mean probability of VTE predicted by the CCM was 0.76%, which would make 13.4% of patients high-risk. The median predicted probability was 0.52% and the range was 0.35-6.3%. Because the distribution of probabilities was highly right-skewed (Figure 2), we selected the median (0.52%) as the primary threshold, which would make 30.6% of patients high-risk.

**Figure 2.**
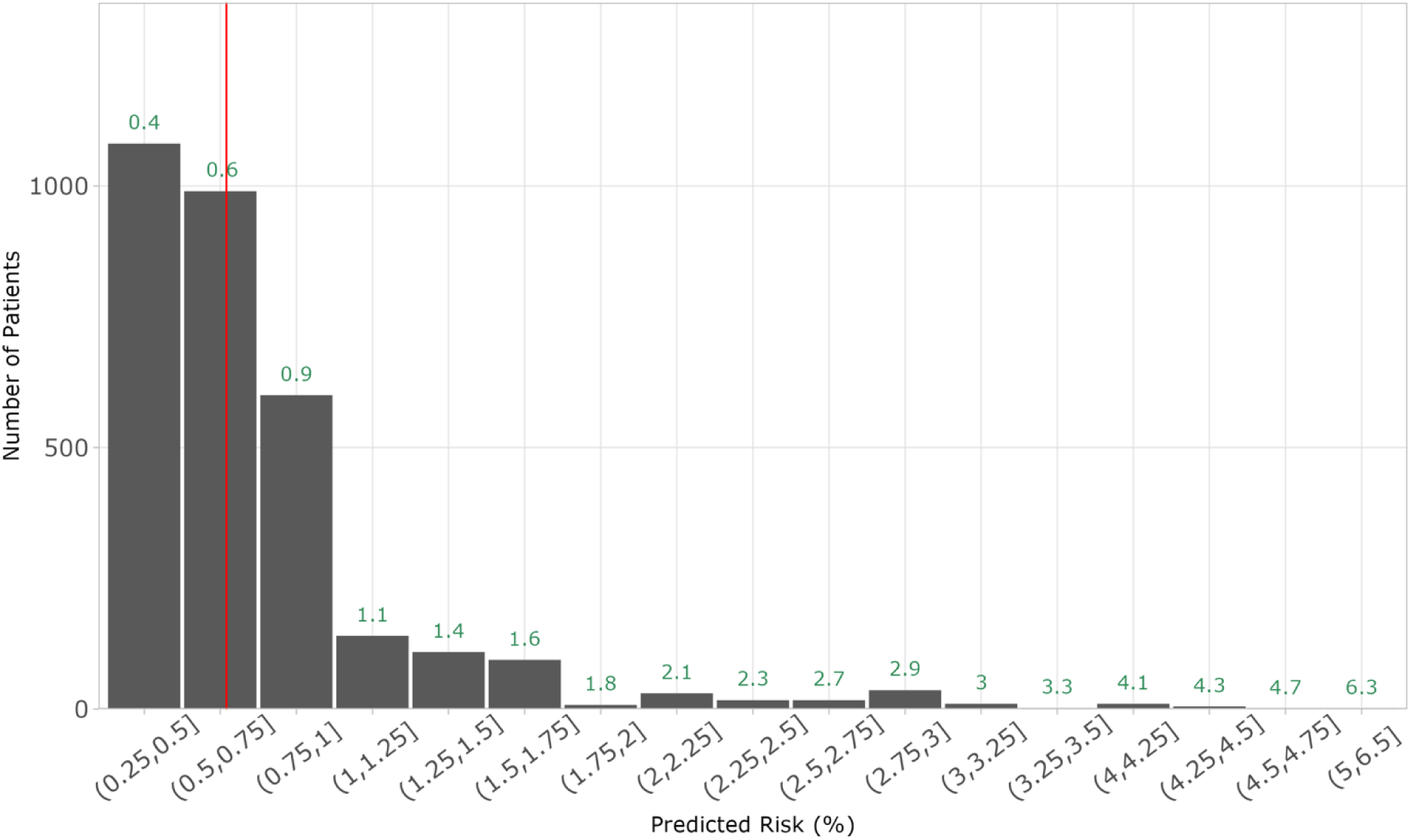
Histogram of risk scores calculated by CCM for patients with a Padua score of 4. The red vertical line indicates the median risk of 0.52%. There is a wide distribution of predicted probabilities that correspond with a Padua score of 4.

### Approach 4: Survey Physicians

In response to the first survey question (Q1), the median physician answered that they would consider a probability of 5% to be high-risk; the mean of all responses was 5.3% and the range was 1-10%. In response to Q2, the mean NNT was 86.6, the median was 50, and the range was 1-1,000 after exclusion of the two outliers. Due to the skewness of responses, we chose the median of the responses to calculate physicians’ stated threshold, which was 5.4%. Despite the similarity of the thresholds derived from Q1 and Q2, a Pearson’s correlation test showed no significant relationship between individual physicians’ thresholds based on responses to Q1 and Q2 (r = -0.05 [95% CI -0.18, 0.09], p = 0.51; Figure 3). Using either of these thresholds, less than 0.5% of patients would be considered high-risk.

**Figure 3.**
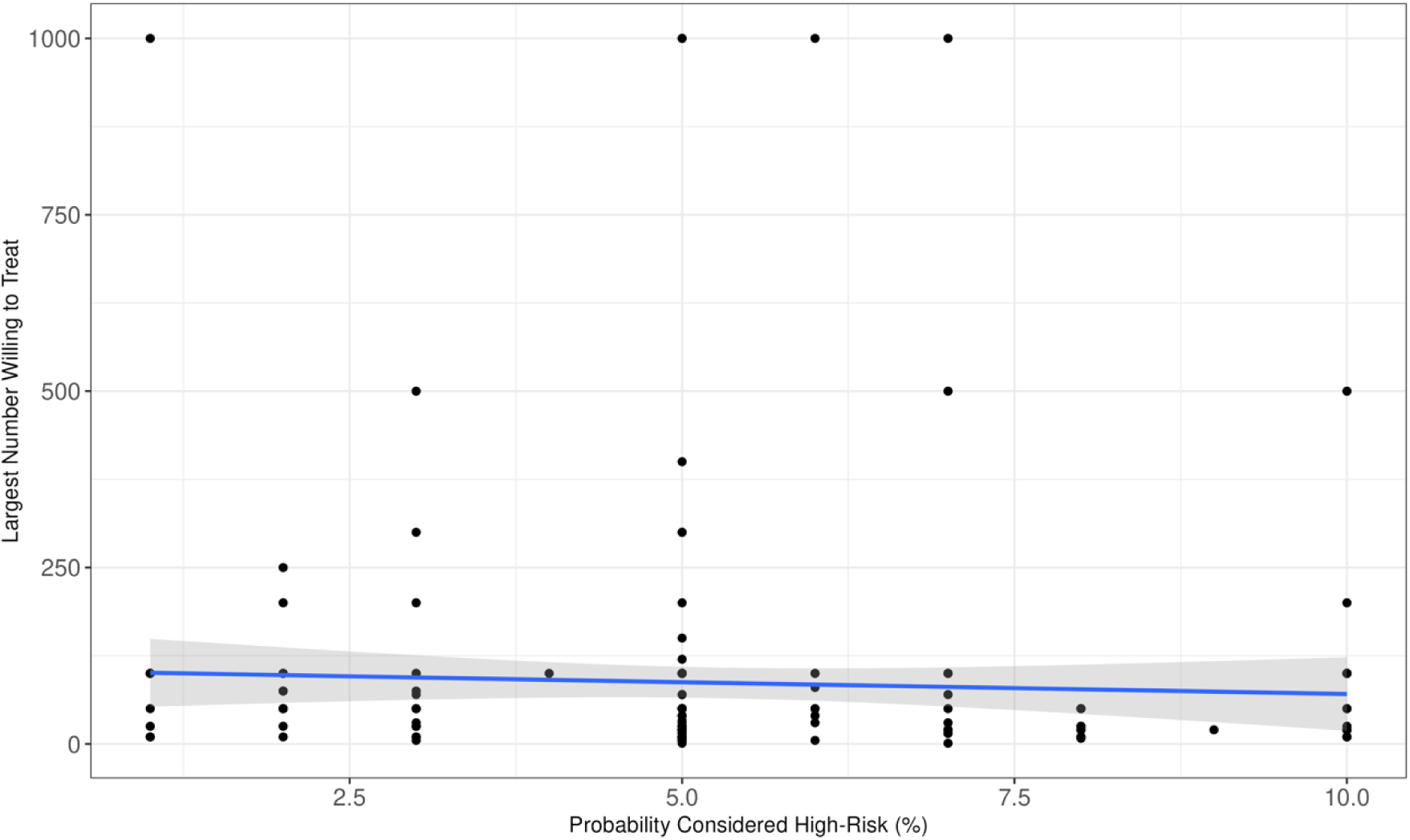
Physician’s responses to Q1 and Q2 of the survey, which asked what probability of developing VTE they would consider high-risk (Q1) and the largest number of patients they would be willing to give prophylaxis to prevent one VTE event (Q2). There was no significant relationship between physicians’ responses to the two analogous questions.

### Approach 5: Examine Physician Behavior

Patients whose physicians did not use the CCM had a high prophylaxis rate (76%). When physicians used the CCM as directed, we observed a lower overall prophylaxis rate (48%) and a smooth gradient in prophylaxis rate, with patients at higher risk prescribed prophylaxis more frequently than those at lower risk (Figure 4). Prophylaxis rates plateaued above the threshold (0.75%) at which patients were designated “high-risk” by the decision support tool. The physicians’ threshold implied by the logistic regression model was 0.55%, which would make 29.2% of patients high-risk.

**Figure 4.**
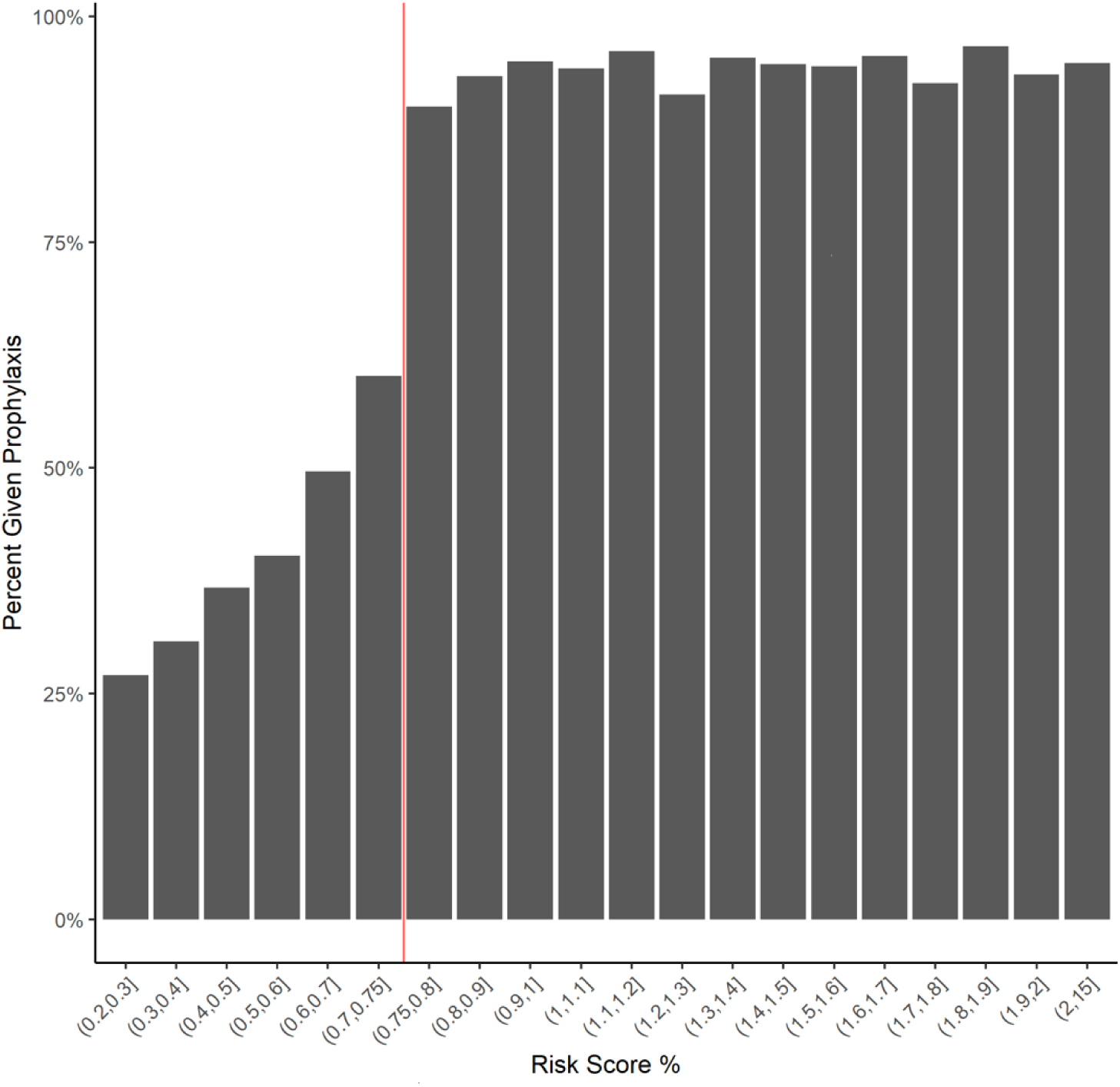
VTE prophylaxis rates for patients who received care guided by the embedded risk calculator, stratified by risk calculated using the CCM. Prophylaxis rates increased with risk, plateauing above the threshold (0.75%; vertical red line) that was provided during the calculator trial period.

## Discussion

In this study, we compared five approaches to deriving an appropriate probability threshold to define “high-risk” for VTE and to guide prophylaxis in medical inpatients. We compared the resulting thresholds in terms of the percentage of patients who would be considered high-risk and thus be recommended to receive prophylaxis. We found that thresholds varied more than tenfold across approaches with corresponding percentages of patients identified as high-risk varying more than 100-fold. The inverse exponential relationship between threshold and patients at high risk suggests that small changes in the threshold can have a substantial impact on the percentage of patients deemed high-risk, particularly within the most “active” decision-making area of the curve (indicated by the shaded box in Figure 1). Probability thresholds within this range could inform the boundaries for desired clinical practice.

The upper limit of this range, a threshold of 1%, would result in 7% of patients being considered high-risk. Although physicians, when asked directly, endorsed an even higher threshold, it is unlikely that hospitals or physicians would be comfortable with such a low rate of prophylaxis eligibility, especially after decades of quality improvement initiatives and hospital quality measures that favor prophylaxis.^22–25^ Moreover, physicians’ observed rates of prophylaxis suggested a much lower threshold. We observed multiple thresholds within a tighter range that reflect a more practical balance between current hospital practices and efforts to curb prophylaxis overuse.^26–29^ Physicians in our cohort who were guided by clinical decision support (CDS) with a threshold of 0.75% displayed equipoise (treating 50% of patients) at a predicted probability of 0.55%. This was only slightly higher than the value (0.52%) that maximized the sum of sensitivity and specificity for that model, which in turn was identical to the median probability of a patient with a Padua score of four. These three approaches offer thresholds ranging from 0.52% to 0.55%, which would result in approximately 30% of medical patients being considered high-risk.

In contrast to this narrow range, the more extreme thresholds we discovered are unlikely to be accepted in clinical practice. The lowest potential threshold (0.3%), based on the cost-indifferent decision analytic model, would lead to about 75% of all patients being considered high-risk. At the other end of the spectrum, based on what physicians said they considered high-risk (>5%), less than 0.5% of patients would be labeled high-risk. Physicians’ stated threshold is clearly impractical, and we observed substantial discrepancy between what physicians say they consider to be high-risk and their revealed probability thresholds based on their actions. Their observed prophylaxis rate of 76% suggests a probability threshold slightly lower than the cost-indifferent decision model, which yielded the lowest threshold of any method we examined. Even when physicians used CDS to identify high-risk patients, they still prescribed prophylaxis to almost half their patients.

Why do physicians’ beliefs about VTE risk and prophylaxis diverge so much from their practices? One possibility is a lack of understanding of probabilities.^30–34^ Rather than viewing risk as a continuous variable, some treat it as a categorical marker.^35^ According to this thinking, patients with risk factors are considered high-risk, whereas those without identified risk factors are not. Indeed, most guidelines approach risk in this way.^36–40^ Even when numbers are used, they generally refer to abstract scores existing on arbitrary scales rather than probabilities. This semi-quantitative approach hinders comparison across risk models and the ability to decide on thresholds for new models. Defining “high-risk” using a probability threshold could help solve these problems, standardize practice across clinicians and hospitals, and improve risk communication and shared decision-making between physicians and patients.^41,42^ Furthermore, meaningful use of CDS for VTE prophylaxis (including standardization of probability thresholds) can mitigate the influence of physician preferences and beliefs about risk on prescribing.

Standardization is important, but for VTE prophylaxis and many other conditions, the appropriate threshold will vary by practice setting. Factors driving this variation may include cost-effectiveness, resource availability, local risk factor distributions, and patient preferences. Even so, guideline committees should establish ranges of acceptable thresholds. Our findings suggest that for VTE, the probability threshold could reasonably vary from 0.5% to 1% (depending on the hospital), resulting in prophylaxis rates ranging from 7% to 31%. This is substantially lower than the average prophylaxis rate observed in our sample, which was 48% when physicians consulted CDS and 76% when they did not. Prophylaxis rates vary even more across hospitals nationally,^43,44^ with some reporting rates approaching 100%.^45^ Evidence-based threshold selection informed by population health goals could enhance the efficacy of CDS tools, improving patient outcomes and overall quality of care.

The current study should be viewed within the context of its limitations, which include that our physician and patient samples were obtained from a single health system. But the percentage of patients determined to be high-risk falls within ranges reported by others using a variety of tools.^46^ We used a limited number of methods to determine potential thresholds; other methods could be used.^47^ Future work might try to empirically determine which approaches are best for meeting pre-defined hospital outcome targets or population health goals. This study also has several strengths, including the use of large survey and electronic health record samples, consideration of multiple approaches, and the ability to compare prophylaxis eligibility rates with observed prophylaxis rates across multiple thresholds. Comparisons between risk stratification recommendations for prophylaxis and observed prophylaxis rates are especially important because they highlight the need for specific guidelines to reduce overuse and target high-risk patients in tandem with the best available risk assessment tools.

## Conclusion

Guidelines recommend prescribing pharmacological prophylaxis to medical patients only if they are at high risk for VTE, but there is no consensus definition of high risk. Moreover, risk is often considered a category rather than a continuum along which a threshold should be applied. We compared five approaches to defining a risk threshold and calculated the corresponding proportion of patients who would be considered high-risk. Thresholds varied substantially across approaches, but most clustered in a VTE risk around 0.5%. Standardizing the definition of high-risk could result in more uniform and appropriate patient-centered care for VTE prevention.

## Data Availability

De-identified data may be made available upon reasonable request.

